# Next-Day Serum Glial Fibrillary Acidic Protein Levels to Aid Diagnosis of Sport-Related Concussion

**DOI:** 10.1101/2024.07.31.24310616

**Authors:** William T. O’Brien, James W. Hickey, Steven Mutimer, Lauren J. Evans, Blake D. Colman, Becca Xie, Lauren P. Giesler, Brendan P. Major, Biswadev Mitra, Gershon Spitz, Terence J. O’Brien, Sandy R. Shultz, Stuart J. McDonald

**Author notes:** **Corresponding Author:** Stuart J. McDonald, Department of Neuroscience, School of Translational Medicine, Monash University, 99 Commercial Road, Melbourne, VIC, 3004, Australia.

## Abstract

The diagnostic utility of blood glial fibrillary acidic protein (GFAP) in sport-related concussion (SRC) is unclear. This study measured serum GFAP at either 16-24 hours (h), 24-32h, or 36-52h post-SRC in 156 Australian football players and compared levels with 98 control players without SRC. Median GFAP levels were higher in SRC cases at 16-24h (124.7 pg/mL; p<0.001) and 24-32h (96.2 pg/mL; p<0.001) compared to controls (66.0 pg/mL), but not at 36-52h (62.8 pg/mL). GFAP had an area under the curve of 0.83 at 16-24h and 0.72 at 24-32h. Serum GFAP at 16-24h can be a useful aid in SRC diagnosis.

## Introduction

Diagnosis of sport-related concussion (SRC) is challenging due to the typical delay in expert review and reliance on potentially biased and incomplete athlete-reported symptoms, risking incorrect medical clearance. Objective information is crucial for accurate diagnosis and proper management of SRC.

Astrocytic injury marker glial fibrillary acidic protein (GFAP) has emerged as a leading blood-based biomarker for traumatic brain injury (TBI),^1^ including in an FDA-approved test to rule out the need for a computed tomography (CT) scan.^2,3^ Promisingly, blood GFAP levels also show excellent ability to discriminate CT-negative mild TBI from healthy controls,^4,5^ indicating potential use for aiding the diagnosis of concussion. Nonetheless, findings from emergency department (ED) mild TBI presentations may not reflect the SRC spectrum.

Elevations in blood GFAP levels have been reported after SRC. However, studies to date have found minimal or moderate accuracy in distinguishing collision sport athletes with and without SRC.^6-9^ For instance, the CARE Consortium reported an area under the curve (AUC) value of 0.68 acutely (median 3 hours [h]) and 0.57 at 24-48h (median 41h).^7^ A potential reason for this limited performance is that sample collection timing may have been primarily outside the peak of GFAP levels, with pharmacokinetic modelling of the CARE SRC data set,^10^ and evidence from ED mild TBI,^11^ indicating GFAP levels may peak 12-36h post-injury.

Given this, the aim of this study was to assess the diagnostic utility of serum GFAP in a community-recruited cohort of SRC, specifically evaluating samples collected at or around 24h post-injury and comparing levels to collision control athletes.

## Methods

### Recruitment

Participants were recruited into studies approved by the Monash University Human Research Ethics Committee or The Alfred Health Human Research Ethics Committee. Between April 10, 2021, and June 30, 2024, adult male and female amateur Australian football players with suspected SRC were identified by club medical doctors, physiotherapists or sports trainers using the Concussion Recognition Tool 5/6 or Sport Concussion Assessment Tool (CRT5/6 or SCAT5/6). Controls were recruited through convenience sampling and randomization (age and sex matched) and included a mixture of healthy (un-injured) control players (HC) and players with musculoskeletal injury (MSK). Inclusion/exclusion criteria are as previously described,^12^ except The Alfred study participants (subset of 2024 recruitment) were not excluded for reporting a concussion within the prior six months.

### Data Collection

Testing occurred 24h post-injury/match (range 16-52h), with venous blood and symptom collection protocols previously described.^12^ Serum GFAP levels were quantified with Simoa Neuro 2-plex B or Simoa Neuro 4-plex B assays on a Simoa HD-X Analyzer, with control and SRC samples balanced within each plate. Overall duplicate coefficient of variation was 6.85%.

### Analysis

Statistical analyses were conducted from July 1-16, 2024. Pearson’s chi-squared, Wilcoxon rank sum, and Fisher’s exact tests were used to compare demographics. Spearman’s correlations were used to assess associations between time post-match/SRC and GFAP levels. Kruskal-Wallis test with Dunn’s multiple comparisons groups were used to compare GFAP levels between control participants (combined HC and MSK group; single combined time-bin) and SRC participants at time-bins of 16-24h, 24.01-32h and 36-52h. *pROC* package was used to calculate AUC values comparing controls versus each individual SRC time-bin. Delong’s test was used to compare AUC values for GFAP versus age and BMI adjusted GFAP, and GFAP + SCAT versus SCAT alone. Analyses were two-tailed (p<0.05) and performed using R Studio, version 4.2.2.

## Results

A total of 315 cases of suspected SRC were reported to the research team, with 156 eligible cases opting to participate. A subset of data (from 75 SRC and 56 controls) are published previously.^12^ Controls comprised 84 HC and 14 MSK cases. Sixteen athletes participated in the study on two occasions, and three participated on three occasions, primarily across different years and groups (**Supplementary Table 1**). Six athletes reported a prior concussion within the last six months (**Supplementary Table 2**). Sensitivity analyses showed that the results were highly comparable with or without these participants included, both when considering repeated participation and prior concussion within the previous six months (**Supplementary Tables 3 and 4**). GFAP levels were highly similar between the HC and MSK groups, supporting their combination into a single control group for analysis (**Supplementary Table 4**). Accordingly, final analysis included 98 control and 156 SRC cases, with negligible differences in age and collection time **(Table 1)**.

**Table 1.** Participant Demographic, Sampling Time and SRC Injury Characteristics.

| Characteristic | SRC, n = 156<sup>†</sup> | Control, n = 98<sup>†</sup> | p-value<sup>‡</sup> |
| --- | --- | --- | --- |
| **Sex** |  |  | 0.92 |
| *Male* | 133 (85%) | 84 (86%) |  |
| *Female* | 23 (15%) | 14 (14%) |  |
| **Age (years)<sup>^</sup>** | 22.5 (20.7, 25.9) | 24.2 (21.7, 26.3) | 0.03 |
| **BMI<sup>^</sup>** | 23.7 (22.5, 25.0) | 24.2 (22.9, 25.4) | 0.11 |
| **History of Concussion<sup>^</sup>** |  |  | 0.11 |
| *Yes* | 101 (65%) | 53 (54%) |  |
| *No* | 55 (35%) | 45 (46%) |  |
| **Number of Previous Concussions<sup>^</sup>** | 1.0 (0.0, 3.0) | 1.0 (0.0, 2.0) | 0.06 |
| **Years of Collision Sport<sup>^</sup>** | 13.0 (10.0, 15.0) | 14.0 (10.0, 17.0) | 0.32 |
| **Time post-SRC/match (hours)** | 25.0 (22.2, 29.4) | 23.1 (20.3, 26.0) | 0.002 |
| **Loss of Consciousness<sup>^</sup>** |  |  | NA |
| *Yes* | 42 (27%) | - |  |
| *No* | 105 (67%) | - |  |
| *Unsure* | 9 (6%) | - |  |
| **Memory Impairment<sup>^</sup>** |  |  | NA |
| *Yes* | 56 (36%) | - |  |
| *No* | 100 (64%) | - |  |

<sup>†</sup> n (%); Median (Interquartile Range).
<sup>‡</sup> Pearson's chi-squared test; Wilcoxon rank sum test; Fisher's exact test.
<sup>^</sup> Self-reported.
Abbreviations: SRC, sport-related concussion; BMI, body mass index.

Serum GFAP levels were compared over the 16-52h sampling window. No effect of time was found for control participants (**Figure 1A**), justifying a single control bin for AUC analysis. GFAP levels decreased over time for SRC participants (Spearman r=-0.27 [95% CI -0.41--0.11], p<0.001; **Figure 1B**).

**Figure 1.**
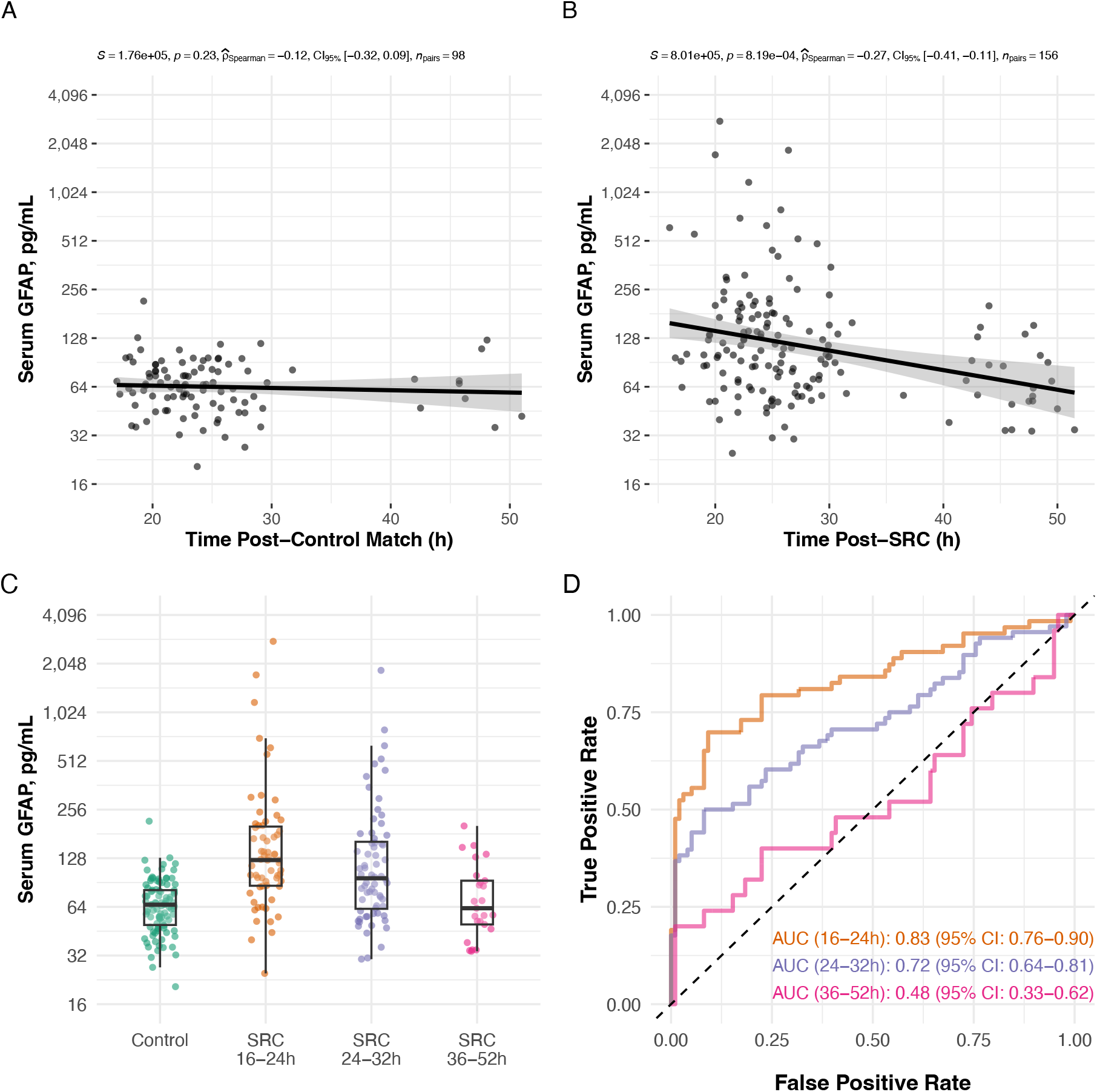
Serum GFAP concentrations and diagnostic performance for SRC over time. A) No association was found between time post-match and GFAP control athletes. B) In contrast, time to sample post-SRC was negatively associated with GFAP levels. C) When separated into SRC time-bins of 16-24h (i.e., early next day; n=63), 24-32h (i.e., late next day; n=68), and 36-52h (i.e. subsequent day; n=25), GFAP levels were elevated at 16-24 and 24-32h compared with control participants. D) Classification accuracy of GFAP for SRC was 0.83 at 16-24h and 0.72 at 24-32h.

When compared with control participants (GFAP median 66.0 pg/mL [IQR, 48.9-81.4]), GFAP levels in SRC participants were higher at 16-24h (median 124.7 pg/mL [IQR, 86.1-203.9]; mean rank difference=82.6, p<0.001) and 24-32h (median 96.2 pg/mL [IQR, 61.0-166.9]; mean rank difference=57.7, p<0.001), but not at 36-52h (median 62.8 pg/mL [IQR, 48.3-96.5]) (**Figure 1C**).

In distinguishing between control and SRC participants GFAP showed an AUC of 0.84 (95% CI, 0.77-0.90) at 16-24h and 0.72 (95% CI, 0.64-0.81) at 24-32h (**Figure 1D**). At 16-24h, a cut-off of 84.5 pg/mL yielded a sensitivity of 0.79 (95% CI, 0.68-0.88) and specificity of 0.78 (95% CI, 0.68-0.85).

Overall GFAP (16-52h) AUC was 0.74 (95% CI, 0.68-0.80) and for next day only (16-32h) was 0.78 (95% CI, 0.71-0.84; **Table 2**). Controlling for age and BMI did not alter GFAP performance. SCAT symptom severity differentiated between groups; however, incremental increases were found when adding GFAP to this measure at 16-24h (AUC=0.92 increased to 0.98, z=2.53, p=0.011).

**Table 2.** Area under the curve statistics for serum GFAP and SCAT symptom severity.

| Time Window | SRC Sample Size | GFAP Raw | GFAP Adjusted | SCAT Symptoms | SCAT Symptoms + GFAP (Raw) |
| --- | --- | --- | --- | --- | --- |
| Next or Subsequent Day | 156 | 0.74<br>(0.68-0.80) | 0.74<br>(0.68-0.80) | 0.93<br>(0.89-0.96) | 0.95<sup>a</sup><br>(0.93-0.98) |
| Next Day | 131 | 0.78<br>(0.72-0.84) | 0.78<br>(0.73-0.84) | 0.93<br>(0.90-0.96) | 0.96<sup>b</sup><br>(0.94-0.98) |
| 16-24 hours | 63 | 0.83<br>(0.76-0.90) | 0.83<br>(0.76-0.90) | 0.92<br>(0.87-0.97) | 0.98<sup>c</sup><br>(0.95-1.00) |
| 24-32 hours | 68 | 0.72<br>(0.64-0.81) | 0.76<br>(0.68-0.83) | 0.93<br>(0.89-0.97) | 0.95<br>(0.91-0.98) |
| 36-52 hours | 25 | 0.48<br>(0.33-0.62) | 0.50<br>(0.35-0.66) | 0.92<br>(0.86-0.99) | 0.93<br>(0.87-1.00) |

Next day defined as 16-32h, subsequent day 36-52h. Data presented as AUC (95% CI). Adjusted indicates GFAP AUC adjusted for age and BMI. Five Control participants and one SRC participant had no SCAT symptom data and were not included in associated AUC calculations.
<sup>a</sup> $z=2.28$ , $p=0.02$ for improvement over SCAT symptoms alone.
<sup>b</sup> $z=2.16$ , $p=0.03$ for improvement over SCAT symptoms alone.
<sup>c</sup> $z=2.53$ , $p=0.01$ for improvement over SCAT symptoms alone.
Abbreviations: SRC, sport-related concussion; GFAP, glial fibrillary acidic protein; SCAT, sport concussion assessment tool. AUC, area under curve; 95% CI, 95% confidence interval; BMI, body mass index.

## Discussion

In our cohort of adult Australian football players with and without SRC, we demonstrate that next-day serum GFAP levels can significantly assist in SRC diagnosis. We found that the timing of GFAP measurement is critical, with a negative correlation between GFAP levels and time to sample over a two-day window after SRC. Within the next-day period (i.e., 16-32h), the AUC values were higher at 16-24h (0.84) than 24-32h (0.72). No utility was found the subsequent day (36-52h). Including GFAP measurements at 16-24h significantly enhanced discrimination between athletes with and without SRC compared to SCAT symptom severity alone, achieving near-perfect separation.

Our findings suggest that previous SRC studies may have underestimated the diagnostic potential of blood GFAP,^6-10^ possibly due to sampling before or after peak concentrations. While diagnostic thresholds and recruitment differences may also play a role, we note that our study involved sideline identification of suspected SRC using CRT/SCAT tools, aligning with the consensus statement on concussion in sport definition of SRC, and included many athletes who would not meet consensus criteria for mild TBI.^13^

While measures at 16-24h post-SRC may represent a biologically optimal time for blood GFAP measures, this period could also serve as a more practical clinic assessment time compared to day-of-injury visits, especially for community sports athletes without “red flag” features for a more serious brain injury.^14^ Moreover, GFAP providing incremental value above SCAT symptoms alone suggests that combining the two may enhance diagnostic certainty. This is significant, given our study participants’ potential for more candid symptom reporting in a research context compared to clinical settings where athletes may be seeking medical clearance. Ultimately, it is essential to transition from relying solely on self-reporting to implementing a new protocol. This protocol should ensure that clinicians routinely receive match-day injury reports, incident footage when available, and objective blood measures of GFAP.

Limitations of this study include the relatively small percentage of female participants, the absence of a requirement for a medical doctor’s diagnosis of SRC, and use of a blood assay that is not yet approved for diagnostic use. Future studies are needed to extend these findings to other cohorts and to generate the evidence required for regulatory approval.

## Conclusion

The findings from this cohort of Australian football players highlight the high accuracy of next-day serum GFAP measurements in distinguishing between collision sport athletes with and without SRC, particularly within 16-24h post-injury. This underscores the potential of next-day serum GFAP as a valuable and objective tool for the early assessment and management of potential or suspected concussion.

## Data Availability

All data produced in the present study are available upon reasonable request to the authors

## References

1. Abdelhak A, Foschi M, Abu-Rumeileh S, et al. Blood GFAP as an emerging biomarker in brain and spinal cord disorders. Nat Rev Neurol. Mar 2022;18(3):158–172. doi:10.1038/s41582-021-00616-3

2. U.S. Food and Drug Administration. (2018) FDA authorizes marketing of first blood test to aid in the evaluation of concussion in adults. https://www.fda.gov/news-events/press-announcements/fda-authorizes-marketing-first-blood-test-aid-evaluation-concussion-adults

3. Bazarian JJ, Biberthaler P, Welch RD, et al. Serum GFAP and UCH-L1 for prediction of absence of intracranial injuries on head CT (ALERT-TBI): a multicentre observational study. Lancet Neurol. Sep 2018;17(9):782–789. doi:10.1016/S1474-4422(18)30231-X

4. Reyes J, Spitz G, Major BP, et al. Utility of Acute and Subacute Blood Biomarkers to Assist Diagnosis in CT-Negative Isolated Mild Traumatic Brain Injury. Neurology. Nov 14 2023;101(20):e1992–e2004. doi:10.1212/WNL.0000000000207881

5. Gill J, Latour L, Diaz-Arrastia R, et al. Glial fibrillary acidic protein elevations relate to neuroimaging abnormalities after mild TBI. Neurology. Oct 9 2018;91(15):e1385–e1389. doi:10.1212/WNL.0000000000006321

6. Meier TB, Huber DL, Bohorquez-Montoya L, et al. A Prospective Study of Acute Blood-Based Biomarkers for Sport-Related Concussion. Ann Neurol. Jun 2020;87(6):907–920. doi:10.1002/ana.25725

7. McCrea M, Broglio SP, McAllister TW, et al. Association of Blood Biomarkers With Acute Sport-Related Concussion in Collegiate Athletes: Findings From the NCAA and Department of Defense CARE Consortium. JAMA Netw Open. Jan 3 2020;3(1):e1919771. doi:10.1001/jamanetworkopen.2019.19771

8. McDonald SJ, O’Brien WT, Symons GF, et al. Prolonged elevation of serum neurofilament light after concussion in male Australian football players. Biomark Res. Jan 10 2021;9(1):4. doi:10.1186/s40364-020-00256-7

9. McDonald SJ, Piantella S, O’Brien WT, et al. Clinical and Blood Biomarker Trajectories after Concussion: New Insights from a Longitudinal Pilot Study of Professional Flat-Track Jockeys. J Neurotrauma. Jan 2023;40(1-2):52–62. doi:10.1089/neu.2022.0169

10. Hier DB, Obafemi-Ajayi T, Thimgan MS, et al. Blood biomarkers for mild traumatic brain injury: a selective review of unresolved issues. Biomark Res. Sep 16 2021;9(1):70. doi:10.1186/s40364-021-00325-5

11. Papa L, Brophy GM, Welch RD, et al. Time Course and Diagnostic Accuracy of Glial and Neuronal Blood Biomarkers GFAP and UCH-L1 in a Large Cohort of Trauma Patients With and Without Mild Traumatic Brain Injury. JAMA Neurol. May 1 2016;73(5):551–60. doi:10.1001/jamaneurol.2016.0039

12. O’Brien WT, Spitz G, Xie B, et al. Biomarkers of Neurobiologic Recovery in Adults With Sport-Related Concussion. JAMA Netw Open. Jun 3 2024;7(6):e2415983. doi:10.1001/jamanetworkopen.2024.15983

13. Silverberg ND, Iverson GL, members ABISIGMTTF, et al. The American Congress of Rehabilitation Medicine Diagnostic Criteria for Mild Traumatic Brain Injury. Arch Phys Med Rehabil. Aug 2023;104(8):1343–1355. doi:10.1016/j.apmr.2023.03.036

14. Echemendia RJ, Brett BL, Broglio S, et al. Sport concussion assessment tool - 6 (SCAT6). Br J Sports Med. Jun 2023;57(11):622–631. doi:10.1136/bjsports-2023-107036

