## Supplementary Tables for "Next-Day Serum Glial Fibrillary Acidic Protein Levels to Aid Diagnosis of Sport-Related Concussion"

**Supplementary Table 1:** Recruitment group and timing for individuals that participated two or three times.

| Participant | Sample #1 | Sample #2 | Sample #3 |
|---|---|---|---|
| 1 | SRC (2021) | MSK (2022) | |
| 2 | SRC (2021) | SRC (2023) | |
| 3 | MSK (2021) | SRC (2024) | |
| 4 | MSK (2021) | HC (2022) | |
| 5 | SRC (2021) | SRC (2024) | |
| 6 | SRC (2021) | HC (2022) | |
| 7 | HC (2022) | SRC (2022) | |
| 8 | HC (2022) | SRC (2023) | |
| 9 | HC (2022) | SRC (2022) | |
| 10 | HC (2022) | HC (2023) | SRC (2024) |
| 11 | SRC (2022) | SRC (2022) | |
| 12 | SRC (2022) | SRC (2024) | |
| 13 | SRC (2022) | HC (2023) | SRC (2023) |
| 14 | HC (2023) | SRC (2024) | SRC (2024) |
| 15 | HC (2023) | SRC (2024) | |
| 16 | SRC (2023) | SRC (2024) | |
| 17 | HC (2023) | SRC (2024) | |
| 18 | SRC (2023) | SRC (2024) | |
| 19 | HC (2023) | SRC (2024) | |

(year) denotes the recruitment year.
Abbreviations: SRC, sport-related concussion; MSK, musculoskeletal; HC, healthy control

**Supplementary Table 2:** Study participants that self-reported sustaining a concussion in the six months prior to recruitment.

| Participant | Group | Time since previous concussion (self-reported; days) |
|---|---|---|
| 1 | SRC | 29 |
| 2 | SRC | 9 |
| 3 | SRC | 29 |
| 4 | HC | 112 |
| 5 | SRC | 50 |
| 6 | SRC | 84 |

Abbreviations: SRC, sport-related concussion; HC, healthy control.

**Supplementary Table 3:** Area under the curve statistics for serum GFAP and SCAT symptom severity for supplementary analyses.

| Time Window | | SRC Sample Size | GFAP Raw | GFAP Adjusted | SCAT Symptoms | SCAT Symptoms + GFAP (Raw) |
|---|---|---|---|---|---|---|
| **Next or Subsequent Day** | *i)* | 156 | 0.74 (0.68-0.80) | 0.74 (0.68-0.80) | 0.93 (0.89-0.96) | 0.95 (0.93-0.98) |
| | *ii)* | 151 | 0.73 (0.67-0.79) | 0.74 (0.67-0.80) | 0.92 (0.89-0.96) | 0.95 (0.93-0.98) |
| | *iii)* | 139 | 0.73 (0.67-0.80) | 0.74 (0.68-0.80) | 0.93 (0.90-0.97) | 0.96 (0.94-0.99) |
| **Next Day** | *i)* | 131 | 0.78 (0.72-0.84) | 0.78 (0.73-0.84) | 0.93 (0.90-0.96) | 0.96 (0.94-0.98) |
| | *ii)* | 127 | 0.77 (0.71-0.83) | 0.78 (0.72-0.84) | 0.93 (0.89-0.96) | 0.96 (0.93-0.98) |
| | *iii)* | 115 | 0.78 (0.72-0.84) | 0.79 (0.73-0.85) | 0.94 (0.91-0.97) | 0.96 (0.94-0.99) |
| **16-24 hours** | *i)* | 63 | 0.83 (0.76-0.90) | 0.83 (0.76-0.90) | 0.92 (0.87-0.97) | 0.98 (0.95-1.00) |
| | *ii)* | 60 | 0.83 (0.76-0.90) | 0.83 (0.76-0.90) | 0.91 (0.86-0.97) | 0.97 (0.95-1.00) |
| | *iii)* | 50 | 0.83 (0.75-0.91) | 0.83 (0.75-0.91) | 0.94 (0.90-0.99) | 0.99 (0.97-1.00) |
| **24-32 hours** | *i)* | 68 | 0.72 (0.64-0.81) | 0.76 (0.68-0.83) | 0.93 (0.89-0.97) | 0.95 (0.91-0.98) |
| | *ii)* | 67 | 0.72 (0.64-0.80) | 0.75 (0.68-0.83) | 0.93 (0.89-0.97) | 0.94 (0.91-0.98) |
| | *iii)* | 65 | 0.74 (0.65-0.82) | 0.76 (0.69-0.84) | 0.93 (0.89-0.97) | 0.94 (0.90-0.98) |
| **36-52 hours** | *i)* | 25 | 0.48 (0.33-0.62) | 0.50 (0.35-0.66) | 0.92 (0.86-0.99) | 0.93 (0.87-1.00) |
| | *ii)* | 24 | 0.48 (0.33-0.64) | 0.52 (0.36-0.68) | 0.92 (0.85-0.99) | 0.93 (0.87-1.00) |
| | *iii)* | 24 | 0.48 (0.33-0.63) | 0.50 (0.34-0.66) | 0.92 (0.85-0.99) | 0.93 (0.87-1.00) |

Next day defined as 16-32h, subsequent day 36-52h. Data presented as AUC (95% CI). Adjusted indicates GFAP AUC adjusted for age and BMI. i) denotes AUC analysis for all data. ii) denotes analysis with exclusion of study participants that self-reported sustaining a concussion in the 6 months prior to recruitment. iii) denotes analysis with the exclusion of the second (and third) participation of an individual. Five control participants and 1 SRC participant had no SCAT symptom data and were not included in associated AUC calculations.

Abbreviations: SRC, sport-related concussion; GFAP, glial fibrillary acidic protein; SCAT, sport concussion assessment tool. AUC, area under curve; 95% CI, 95% confidence interval; BMI, body mass index.

**Supplementary Table 4:** Distribution of GFAP levels for supplementary analyses.

| | Sample size | GFAP Median (IQR) | Sample size | GFAP Median (IQR) |
|---|---|---|---|---|
| | SRC | | Control | |
| **All data** | 156 | 100.07 (63.32 – 169.22) | 98 | 66.00 (49.46 – 81.23) |
| **Concussion in prior six months excluded** | 151 | 100.00 (62.83 – 166.39) | 97 | 65.87 (49.12 – 81.30) |
| **Repeated participation excluded** | 139 | 98.81 (62.83 – 160.99) | 93 | 66.14 (48.03 – 81.05) |
| | MSK | | HC | |
| **MSK vs HC** | 14 | 70.48 (49.88 – 79.87) | 84 | 64.84 (50.15 - 81.93) |

Abbreviations: SRC, sport-related concussion; GFAP, glial fibrillary acidic protein; MSK, musculoskeletal injury; HC, healthy control; IQR, interquartile range.
